# Cost-utility of two minimally-invasive surgical techniques for operable oropharyngeal cancer: Transoral robotic surgery versus transoral laser microsurgery

**DOI:** 10.1101/2020.12.30.20249018

**Authors:** Enea Parimbelli, Federico Soldati, Lorry Duchoud, Gian Luca Armas, John R. de Almeida, Martina Broglie, Silvana Quaglini, Christian Simon

## Abstract

**Importance:** Transoral robotic surgery (TORS) and transoral laser micro-surgery (TLM) are two different but competing minimally invasive techniques to surgically remove operable oropharyngeal squamous cell cancers (OPSCC). As of now, no comparative analysis as to the cost-utility of these techniques exists.

**Objective:** Recent population-level data suggest for TORS and TLM equivalent tumor control, but different total costs, need for adjuvant chemoradiation, and learning curves. Therefore, the objective of this study was to compare TORS and TLM from the cost-utility (C/U) point of view using a decision-analytical model from a Swiss hospital perspective.

**Design:** Our decision-analytical model combines decision trees and a Markov model to compare TORS and TLM strategies. Model parameters were quantified using available literature, original cost data from two Swiss university tertiary referral centers, and utilities elicited directly from a Swiss population sample using standard gamble. C/U and sensitivity analyses were used to generate results and gauge model robustness.

**Setting:** Swiss hospital perspective

**Intervention:** Cost-utility analysis

**Main outcome measure:** Comparative cost-utility data from TLM and TORS

**Results:** In the base case analysis TLM dominates TORS. This advantage remains robust, even if the costs for TORS would reduce by up to 25%. TORS begins to dominate TLM, if less than 59,7% patients require adjuvant treatment (pTorsAlone>0.407), whereby in an interval between 55%-62% (pTorsAlone 0.38-0.45) cost effectiveness of TORS is sensitive to the prescription of adjuvant CRT. Also, exceeding 29% of TLM patients requiring a re-operation for inadequate margins renders TORS more cost-effective.

**Conclusion:** TLM is more cost-effective than TORS. However, this advantage is sensitive to various parameters i.e. the number of re-operations and adjuvant treatment.

**Key points:** *Question:* Compare cost-utility of TORS versus TLM

*Findings:* In the base case analysis TLM dominates TORS, even if the costs for TORS would reduce by up to 25%. TORS begins to dominate TLM, if less than 59,7% patients require adjuvant treatment, whereby in an interval between 55%-62% cost effectiveness of TORS is sensitive to the prescription of adjuvant CRT. Exceeding 29% of TLM patients requiring a re-operation for inadequate margins renders TORS more cost-effective.

*Meaning:* TLM is more cost-effective than TORS. However, this advantage is sensitive to the number of re-operations and adjuvant treatment

## INTRODUCTION

Transoral Robotic Surgery (TORS) and Transoral Laser Microsurgery (TLM) are two minimally-invasive surgical approaches for the treatment of oropharyngeal cancer. In the past few decades a re-evaluation of treatment paradigms with a desire to spare patients the treatment-related toxicities of chemoradiotherapy, has led to an increased use of new minimally invasive surgical techniques to improve outcomes^1,2^.

TORS is a technique that utilizes wristed robotic surgical technology through a transoral approach in order to facilitate en-bloc resection of tumors. Resections are performed using electrocautery with endoscopes providing different angles of visualization. This technique provides excellent visualization of the disease and possibility to resect the tumor in one piece allowing for more precision in terms of margin analysis^3^.

TLM, on the other hand, utilizes lasers, and visualization through laryngoscopes and mouth gags for exposure. Typically, the field of vision is smaller than with TORS, thus the laryngoscopes have to be repositioned several times during the intervention. TLM follows the philosophy of resecting the tumor in pieces. This may confer a better control over the deep margin as a consequence of traversing the tumor and assessing the different cauterisation characteristics between tumor and normal tissue, it may add however a certain degree of uncertainty to the final reading of the margins^4^.

A recent population-level analysis demonstrated equivalent survival and similar positive margin rates with both techniques, but a significantly higher rate of post-operative chemoradiation in the TLM group^5^ suggesting that uncertainties over margins may lead treating physicians to rather favour the more aggressive postoperative treatment.

TLM appears to have a steeper learning curve than TORS. This is of importance to head and neck cancer programs wanting to implement one or the other technique. To the contrary, TLM infers less total costs as a consequence of the high equipment and disposable costs incurred during robotic^6^.

In summary, a comparative analysis of the two minimally-invasive techniques is warranted for which we used a decision-analytic model for comparing TORS and TLM from the cost-utility (C/U) point of view.

Despite minimally-invasive techniques have been proven to be cost-effective in head&neck cancer^7,8^, to the best of our knowledge, there is no literature directly comparing TORS and TLM.

## MATERIALS AND METHODS

Our base case consists of a Swiss patient with an oropharyngeal squamous cell carcinoma (OPSCC), age 55, with operable T-category (T1 or T2) OPSCC. Our analyses are performed from a Swiss hospital payer perspective and with a lifetime horizon.

We developed a two-stage model based on (i) a published model about the economic evaluation of TORS vs radiotherapy^8^, (ii) additional literature^5^ and (iii) authors’ expertise and statistics from the Centre Hospitalier Universitaire Vaudois (CHUV) and University Hospital Zurich (USZ) hospitals.

The first-stage decision tree accounts for short-term outcomes of the surgery and its complications which are, in turn, carried forward as initial conditions for a second-stage model representing long-term outcomes through a Markov process.

The first-stage model is depicted in figure 1. The two surgical strategies constitute alternatives for the first decision node, after which a chance node distinguishes between cases undergoing surgery alone, and cases requiring adjuvant radiotherapy (RT) or chemoradiotherapy (CRT). Finally, potential complications of the surgical interventions and, where appropriate, associated adjuvant (ADJ) therapy are modeled.

**Figure 1.**
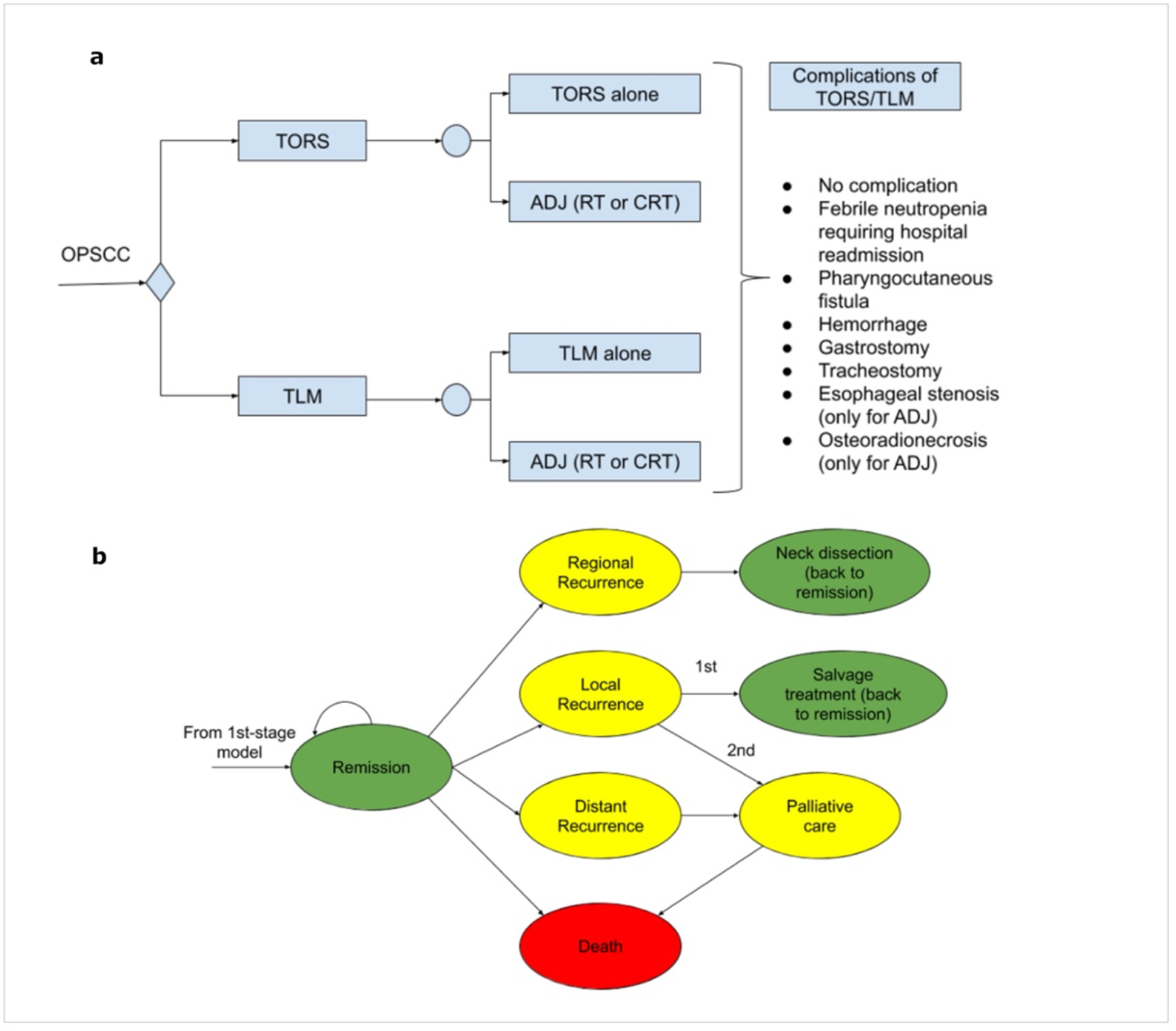
short-term outcomes decision tree (a) and second-stage Markov model (b).

The second-stage model deals with long-term outcomes and is constituted by a Markov model (figure 1b). It represents patients entering a state of remission after treatment and models their transitions through other possible health states until death. The Markov cycle has been set to 3 months and the time horizon is the entire patient life. Initial rewards of each Markov model are carried forward from the results of the first-stage model.

Model parameters representing estimates of probabilities of adjuvant treatment were derived from data published by Li et al.^5^ which constitute the largest and most recent published database study with relevant outcome data. Other parameters modeling clinical events such as complication rates and recurrence rates were determined from systematic review of the literature^9^. The hospital admission rate for CRT was set at 75% of patients to be admitted once, and 25% twice. The proportion of patients needing hospital admission for RT was set at 25% only once. Regarding the need for a gastrostomy we considered a PEG-rate for CRT of 70% while 20% for RT as per institutional data from CHUV and USZ.

Transition probabilities between different health states of the Markov models were directly adopted from de Almeida et al.^8^ and no relevant difference in survival is assumed between the TORS and TLM arms of the model, based on a recent retrospective analysis of the National Cancer Data Base (NCDB)^5^. Risk of death from non-cancer-specific causes is modeled following Swiss life tables, acquired through the Swiss *office federal de la statistique*. In order to perform probabilistic sensitivity analysis (PSA) all parameters were represented using probability distributions. Probabilities of event occurrence were represented as beta distributions, as indicated for variables ranging from 0 to 1^10^ (Table 1).

Costs were directly acquired from the “Centre hospitalier universitaire vaudois (CHUV)” and “Universitätsspital Züerich (USZ)” administrative departments. Having adopted a hospital perspective, costs incurred by the patient are not considered in our analyses. All costs are represented as Gamma distributions, as suggested by Huinink et al^10^ for values greater or equal than 0 (Supplementary??Table 1). Utility coefficients (UCs) for the health states included in the model were collected with Standard Gamble method through our UceWeb^11,12^ platform from a set of 41 Swiss healthy volunteers. 17 different scenarios were evaluated by each participant. Rating Scale method was also administered, to familiarize participants with the tool and as a consistency check of the obtained values. As for probabilities, UCs are represented as beta distributions^7^ (Table1).

Willingness-to-pay was set to 4000 CFH/QALM (i.e. 48000 CFH/QALY^13,14^. Incremental cost was computed from the difference in expected cost (CFH) between TORS and TLM. Similarly, incremental utility was computed from the difference in expected utility between TORS and TLM. The incremental cost-utility ratio was derived taking the quotient between incremental cost and incremental utility. All cost-utility analyses were performed using TreeAge Pro 2019 software (Williamstown, MA, 2019).

Key model parameters were varied using one-way and two-way deterministic sensitivity analysis in order to assess their impact on the results. In particular, we explored the key role of adjuvant therapy (RT or CRT) after surgery and costs of treatment. Furthermore, all parameters affected by uncertainty were varied in probabilistic sensitivity analysis (PSA). Probabilistic sampling was performed from the distributions described above for probabilities (Beta), costs (Gamma) and utilities (Beta). PSA was performed using second-order Monte-Carlo simulations using 1000 simulations. Incremental cost and effectiveness were plotted with 95% confidence ellipsoids.

## RESULTS

The base case analysis used model parameters presented in the methods section and Table 1. It shows that TORS is moderately more effective than TLM (Months: 342.72 versus 342.62) but also more costly than TLM (Costs in CHF 56879.13 versus 53518.28). When taking into account quality of life, TLM dominates TORS with slightly higher QALMs (216.40 versus 216.31) at a lower cost.

An important role is assumed by the adoption of adjuvant (chemo)radiotherapy after TORS and TLM. Univariate sensitivity analyses on effectiveness show that when the probability of adjuvant therapy is less than 1-0.403 (<0.597) for TORS, TORS is the optimal option. The same is true for TLM when the probability of adjuvant therapy is less than 1-0.38 (<0.62). When varied simultaneously in a 2-way sensitivity analysis the effect on the optimal strategy is also evident, as the modality with a lower chance of needing adjuvant treatment is preferred by the model (Figure 2).

**Figure 2.**
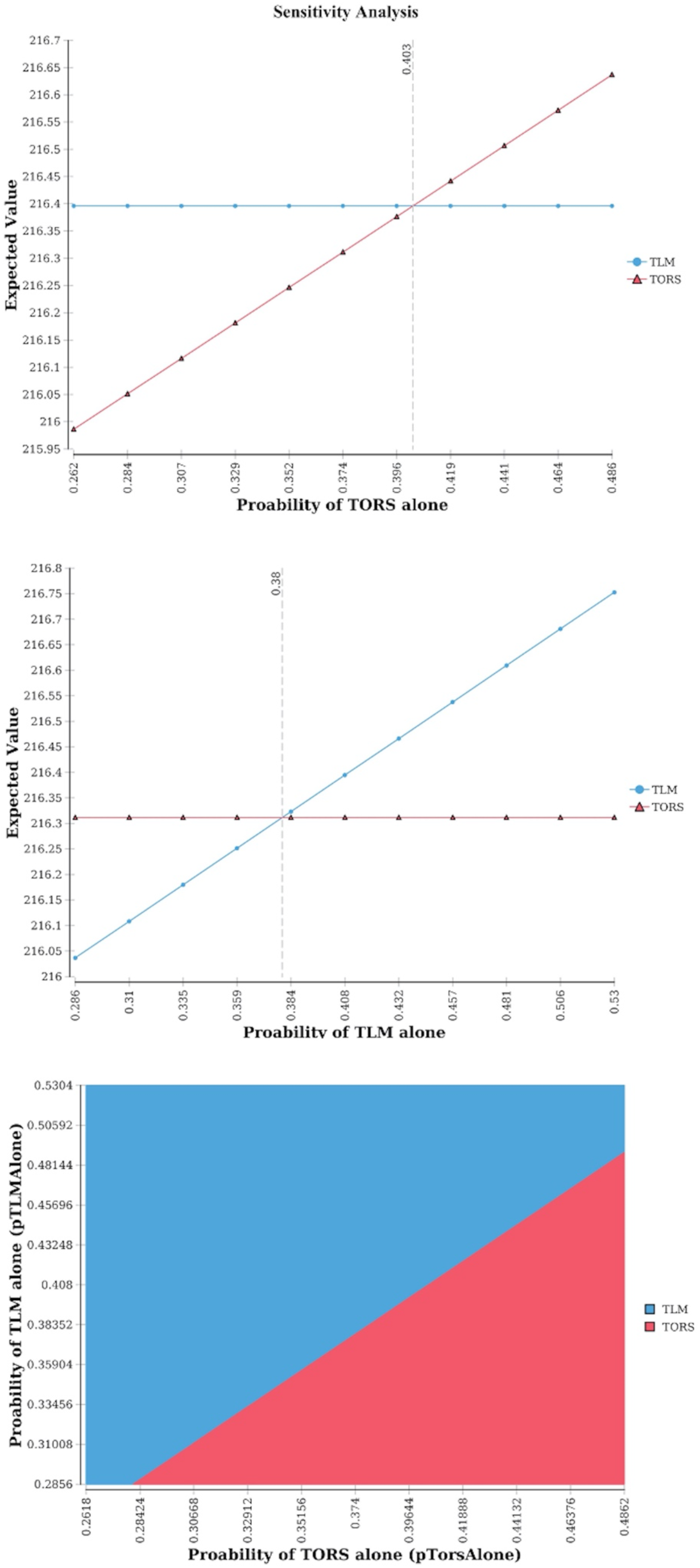
1-way and 2-way sensitivity analyses of probability of TORS alone and probability of TLM alone.

In the hypothetical scenario where improvements to TORS, or careful patient selection, vary the proportion of patients needing adjuvant therapy (i.e. pTorsAlone varies) also the type of the adjuvant treatment begins to play a role. Decreasing the proportion of patients that receive chemoradiotherapy, instead of radiotherapy, as adjuvant treatment after TORS can also make TORS a preferred option over TLM. A 2-way sensitivity analysis on pCRT_Tors and pTorsAlone shows that if the use of adjuvant therapy is lower than 1-0.55 (<0.45), then TORS is the preferred option. However, if the use of adjuvant therapy after TORS is between 0.55 – 0.62 (pTorsAlone is in the [0.38-0.45] interval) the proportion of patients receiving CRT as adjuvant therapy has to stay under a certain value for TORS to be the optimal alternative (Figure 3).

**Figure 3.**
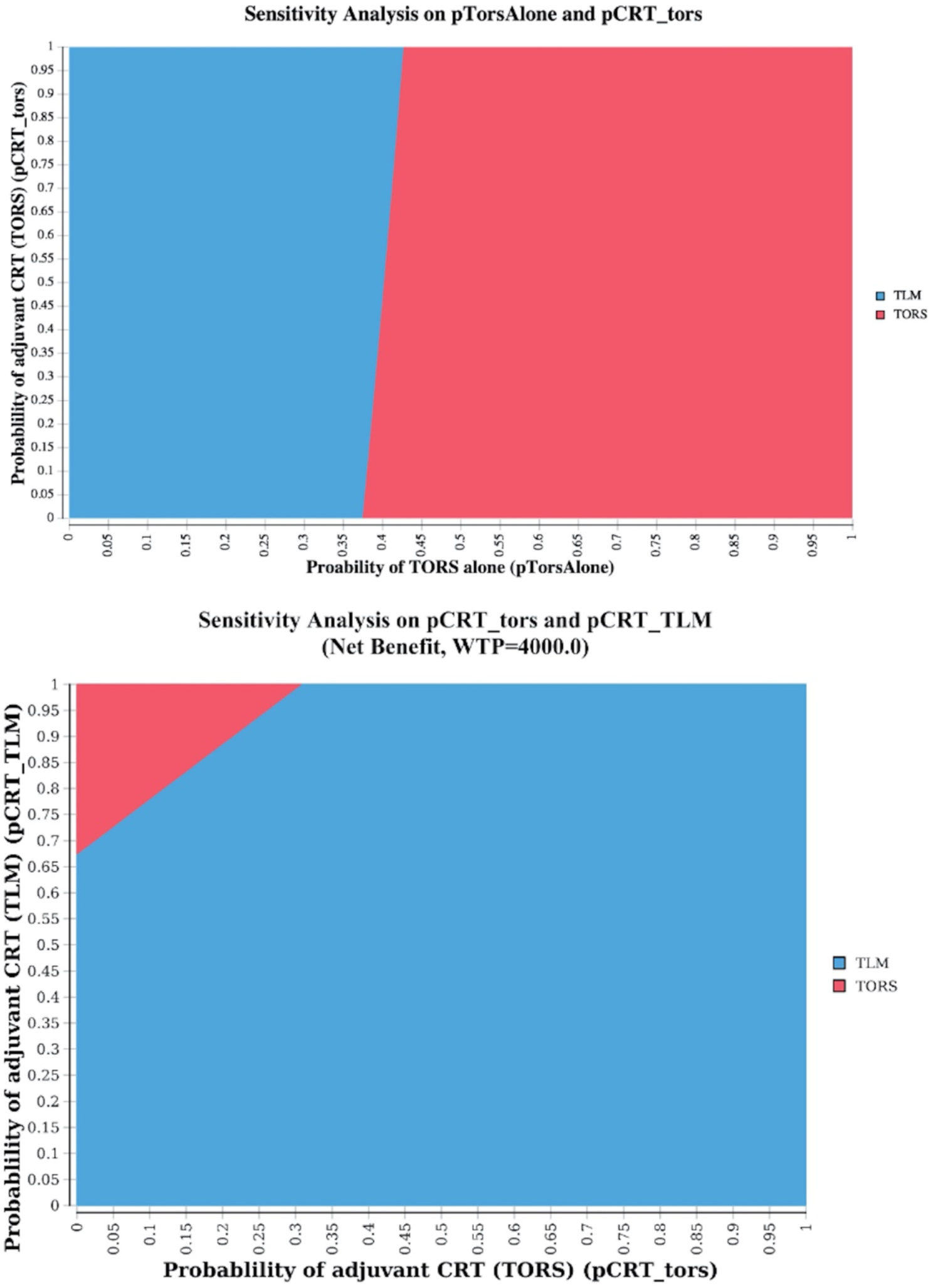
2-way sensitivity analyses for pTorsAlone and pCRT_Tors, and probability of adjuvant chemoradiotherapy after TORS and after TLM.

A 2-way sensitivity analysis (Figure 3) shows how being able to decrease the proportion of patients that receive chemoradiotherapy, instead of radiotherapy alone, can make TORS a preferred option over TLM. It shows how the combination of values of pCRT_TORS and pCRT_TLM that sees TORS the preferred option is rather small.

Given that during TLM the tumor is resected in multiple pieces^15^, it is common in clinical practice to bring back patients for one or multiple resections in order to achieve negative margins. Our analysis shows that as soon as TLM needs to be repeated once in more than 29% of the patients (krepeatTLM >=2, threshold value = 1.292 in sensitivity analysis), the increased cost compared to TORS makes TORS the preferred option with a higher net monetary benefit (Figure 4). On the other hand, results of the base case analysis are rather robust to changes in cost of TORS, confirming TLM as the optimal option even for a relevant (∼24%) decrease in TORS cost. A 2-way sensitivity analysis highlights, how repeating TLM even only once (krepeatTLM=2) and keeping TORS cost as-is, results in TORS being the preferred option for cost-effectiveness (Figure 4).

**Figure 4.**
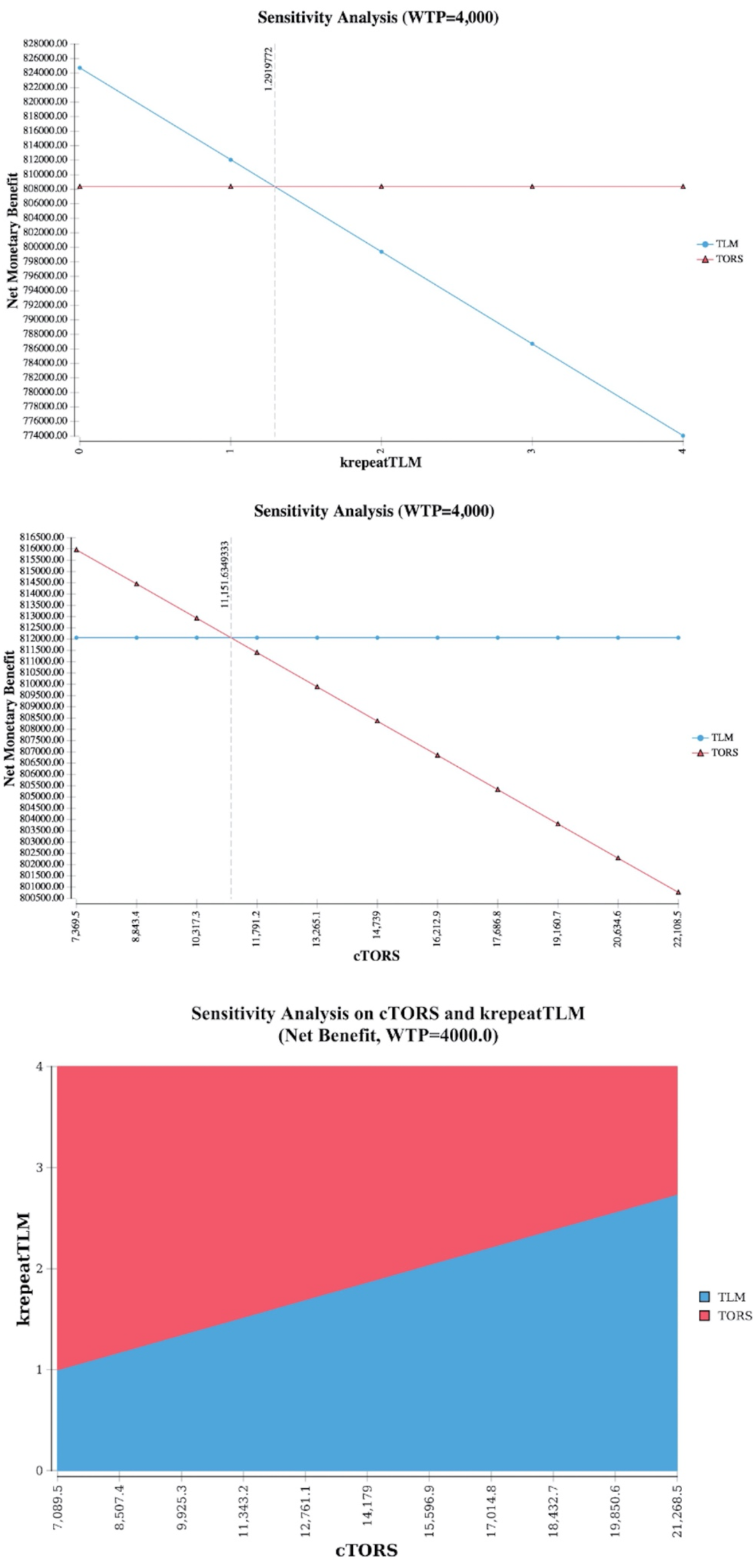
1 way sensitivity analysis on cost of TLM (based on number of re-resections needed for negative margins), cost of TORS, and 2-way sensitivity analysis combining the two. Note: NMB = WTP*QALMs.

Using base-case parameters, PSA shows the majority of simulations favoring TLM, having lower cost (incremental cost is >0 for most simulations) and close to 0 incremental effectiveness compared to TORS. In the 1000 simulations, with a willingness to pay of 4000 CFH/QALM (i.e. 48000 CHF/QALY), TORS dominates in only 1.1% of the cases, is cost-effective in 5.2%, while TLM is cost-effective in 66% of the cases, and dominates in the remaining 27.7% (Figure 5).

**Figure 5.**
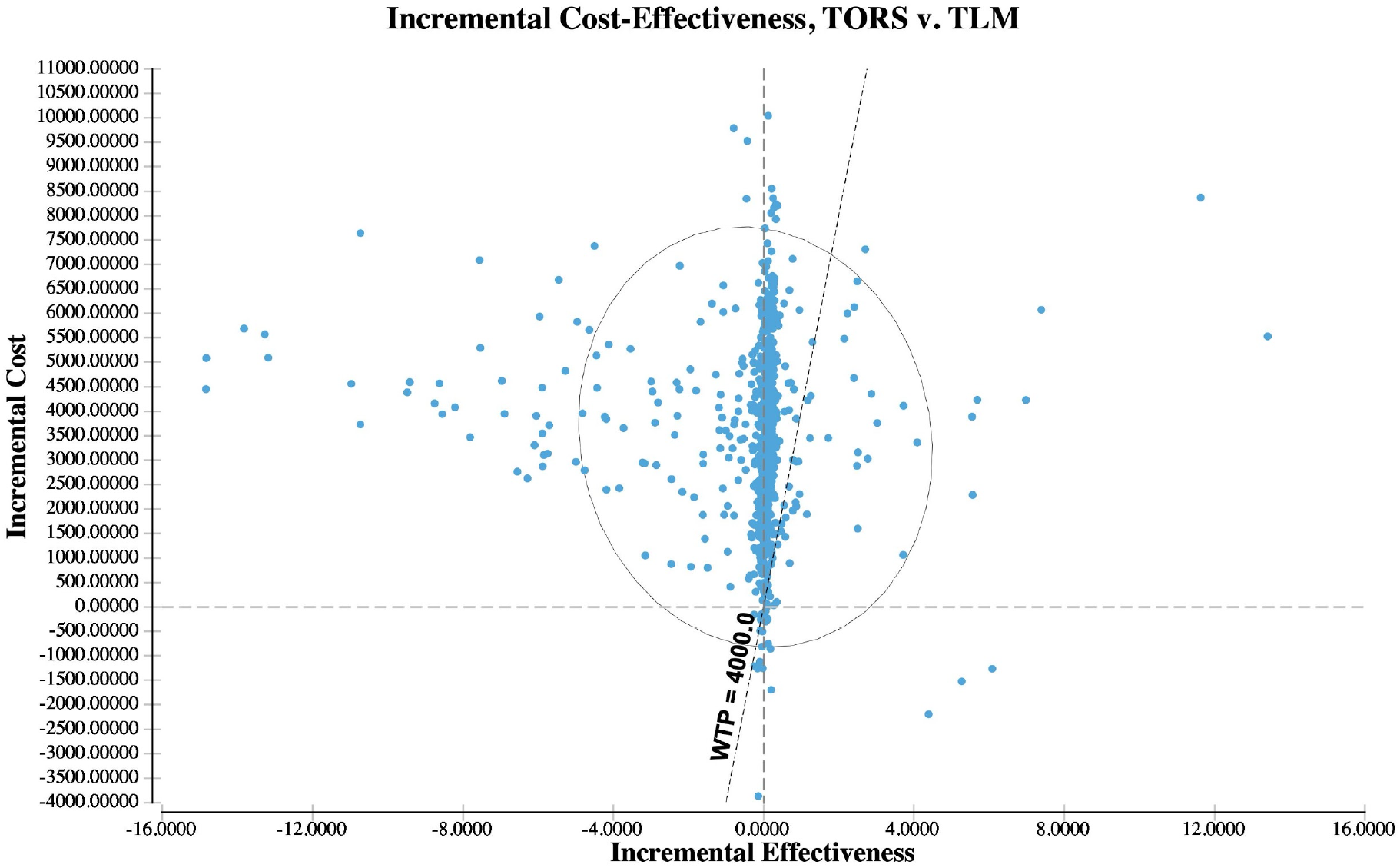
PSA, base case analysis.

## Discussion

Our two-stage model-based analysis shows that TLM is currently a cost-effective surgical treatment choice for operable OPSCC. Results, albeit robust, are sensitive to a variability in the proportion and nature of adjuvant therapy and need of performing TLM re-resections impacting costs.

TORS becomes more cost-effective than TLM, if the rate of post-operative adjuvant treatment is below 59,7% and is sensitive to the replacement of RT by CRT in an interval of a rate of adjuvant treatment between 55% and 62%. Higher rates of postoperative therapy after TORS reduce its overall utility and suggest that a careful case selection, limiting cases needing adjuvant chemoradiotherapy, might be important to impact its cost-effectiveness.

The potential need to repeat TLM surgery for close or positive margins, even only once, results in a significant change in TLM costs, favoring TORS as the cost-effective alternative. According to our one-way sensitivity analysis on costs of TLM (Figure 4) the threshold for TORS to be more cost-effective as TLM based on the number of re-interventions is found to be at 1.29, suggesting that if 29% or more patients require re-resections after TLM and given that none of the TORS patients’ need to be taken back to the OR outside the regular setting used for the base case scenario, TORS is superior in terms of cost-effectiveness. This percentage is already reached in certain centers even with large experience according to current literature. In a study comprising of 1467 patients treated with TLM for cancers of the oral cavity, oropharynx, larynx, and hypopharynx, 386 patients (26,3%) were taken back for a second resection, and of those even another 22 for a third and a fourth resection with TLM^17^. It seems therefore critical to avoid second operations with TLM by eventually relying more on the use of frozen sections whenever and wherever feasible.

In general, advantages of TORS are the learning curve, allowing for easier adaptation of the surgeon and better results in a shorter amount of time^18^. A clear disadvantage are the upfront capital costs that are widely exceeding the costs of TLM^6^. TLM to the contrary has lower upfront costs, but is technically more challenging requiring more training and a longer time until mastering the procedure^16^.

There are certainly limitations of this type of analysis. Modeling is based on various parameter estimates, most of which are retrospectively taken from various sources. Also, this analysis has been performed from a Swiss hospital perspective. It is probable that other healthcare systems account for other costs eventually limiting the generalizability of the results.

The data presented in this study may suggest that TLM is superior with respect to C/U to TORS. However, the decision making on implementing a TORS or TLM program should be based on additional objectives, i.e. the use of a robotic platform for i.e. endoscopic thyroid and neck surgery and/or other applications of the robot. While TLM is based on a technology platform less easy to expand, TORS uses technology for which new applications are easier to identify.

In summary, in this study we provide evidence for an advantage of TLM over TORS in terms of cost-effectiveness for the surgical minimally invasive treatment of operable OPSCCs. However, this advantage is sensitive to the rate of adjuvant treatment, the prescription of RT versus CRT, and the rate of patients requiring re-resections for inadequate margins.

## Data Availability

All relevant data is included in the manuscript and its supplementary material

