## supplemental material for "Cost-utility of two minimally-invasive surgical techniques for operable oropharyngeal cancer: Transoral robotic surgery versus transoral laser microsurgery"

**Table 1** - Model parameters: Probabilities of occurrence of events, costs and utilities.

**NOTE: for prr, plr and pdr 80% of recurrences were modeled in the first 2 years, and the remaining 20% between 2 and 5 years posttreatment (probabilities were adjusted accordingly, assuming 5% of patients have recurrences in the first 2 years[6])*

| Variable name | Description | Mean | Standard deviation | Distribution type | Parameter 1 (alpha) | Parameter 2 (beta) |
|---|---|---|---|---|---|---|
| | | | **Probabilities of events** | | | |
| *pes* | Probability of esophageal stenosis | 0.0476 | 0.0005 | Beta | 4 | 80 |
| *phem* | Probability of hemorrhage | 0.0243 | 0.0001 | Beta | 6 | 241 |
| *pho_adj* | Probability of hospital readmission after adjuvant | 0.1731 | 0.0027 | Beta | 9 | 43 |
| *pho_s* | Probability of hospital readmission (TORS or TLM) | 0.0333 | 0.0010 | Beta | 1 | 29 |
| *plg* | Probability of long-term gastrostomy (1 year) after adjuvant treatment | 0.0500 | 0.0003 | Beta | 9 | 171 |
| *plt* | Probability of long-term tracheostomy (1 year) | 0.0226 | 0.0001 | Beta | 4 | 173 |
| *psg* | Probability of short-term (6 months) gastrostomy (TORS or TLM) | 0.0144 | 0.0001 | Beta | 2 | 137 |
| *psg_adj* | Probability of short-term (6 months) gastrostomy after adjuvant | 0.2991 | 0.0019 | Beta | 32 | 75 |
| *por* | Probability of osteoradionecrosis | 0.0265 | 0.0002 | Beta | 4 | 147 |
| *ppf* | Probability of pharyngocutaneous fistula | 0.0253 | 0.0001 | Beta | 10 | 385 |
| *pTLMAlone* | Probability of TLM alone | 0.4085 | 0.0007 | Beta | 134 | 194 |
| *pTorsAlone* | Probability of TORS alone | 0.3740 | 0.0001 | Beta | 824 | 1379 |
| *pCRT_TLM* | Probability of adjuvant CRT (TLM) | 0.6289 | 0.0012 | Beta | 122 | 72 |
| *pCRT_tors* | Probability of adjuvant CRT (TORS) | 0.5272 | 0.0002 | Beta | 727 | 652 |
| *pRT_TLM* | Probability of adjuvant RT (TLM) | 0.3711 | 0.0012 | Beta | 72 | 122 |
| *pRT_tors* | Probability of adjuvant RT (TORS) | 0.4728 | 0.0002 | Beta | 652 | 727 |
| *plr** | Probability of local or regional recurrence (first 2 years) | 0.0064 | 0.0000 | Beta | 11 | 1715 |
| *prr** | Probability of regional recurrence (first 2 years) | 0.0064 | 0.0000 | Beta | 11 | 1715 |
| *pdr** | *Probability of distant recurrence (first 2 years)* | *0.0038* | *0.0000* | *Beta* | *11* | *2900* |
| **Costs (CHF)** | | | | | | |
| *cTORS* | *Cost of TORS* | *14739* | *869.31* | *Gamma* | *287.4635* | *0.0195* |

Deleted: F

| | | | | | | |
|---|---|---|---|---|---|---|
| *cTLM* | *Cost of TLM* | *12671* | *516.23* | *Gamma* | *602.4698* | *0.0475* |
| *cCRT* | Cost of adjuvant CRT | 33911 | 2079.08 | Gamma | 266.0350 | 0.0078 |
| *cRT* | Cost of adjuvant RT | 27962 | 1714.35 | Gamma | 266.0342 | 0.0095 |
| *cES* | Cost of esophageal stenosis | 2362 | 410.65 | Gamma | 33.0832 | 0.0140 |
| *cGAST* | Cost of gastrostomy | 4332 | 410.65 | Gamma | 111.2820 | 0.0257 |
| *cHR_adj* | Cost of hospital readmission (for adjuvant) | 10097 | 619.05 | Gamma | 266.0342 | 0.0263 |
| *cHR_s* | Cost of hospital readmission (TORS or TLM) | 8203 | 803.41 | Gamma | 104.2498 | 0.0127 |
| *cORN* | Cost of osteoradionecrosis | 32111 | 1077.71 | Gamma | 887.7769 | 0.0276 |
| *cPF* | Cost of pharyngocutaneous fistula | 82892 | 333.96 | Gamma | 61609.3654 | 0.7432 |
| *cPH* | Cost of hemorrhage (from surgical site) | 4469 | 415.50 | Gamma | 115.6865 | 0.0259 |
| *cTRACH* | Cost of tracheostomy | 11688 | 612.67 | Gamma | 363.9366 | 0.0311 |
| *cREM* | Cost of remission 0-2 y | 168.5 | 9.68 | Gamma | 303.2588 | 1.7998 |
| *c2REM* | Cost of remission 2-5 y | 60 | 9.68 | Gamma | 38.4518 | 0.6409 |
| *cPC* | Cost of palliative care | 4137 | 367.86 | Gamma | 126.4754 | 0.0306 |
| *cRR* | Cost of regional recurrence | 7047 | 464.24 | Gamma | 230.4227 | 0.0327 |
| *cLR_chemorad* | Cost of local recurrence (chemoradiation) | 34041 | 2079.08 | Gamma | 268.0786 | 0.0079 |
| *cLR_s* | Cost of local recurrence (surgical resection) | 40513 | 2050.27 | Gamma | 390.4511 | 0.0096 |
| *cDM* | Cost of distant metastasis | 4137 | 367.86 | Gamma | 126.4754 | 0.0306 |
| *cPanendo* | Cost of panendoscopy | 388 | 23.79 | Gamma | 266.0337 | 0.6857 |
| **Utilities** | | | | | | |
| *uSURG* | Utility coefficient of TORS or TLM | 0.902 | 0.203 | Beta | 1.0328 | 0.1122 |
| *uRT* | Utility coefficient of adjuvant RT | 0.850 | 0.275 | Beta | 0.5831 | 0.1029 |
| *uCRT* | Utility coefficient of adjuvant CRT | 0.794 | 0.317 | Beta | 0.4984 | 0.1293 |
| *uHR* | Utility coefficient of hospital readmission | 0.954 | 0.140 | Beta | 1.1820 | 0.0570 |
| *uPF* | Utility coefficient of pharyngocutaneous fistula | 0.932 | 0.194 | Beta | 0.6374 | 0.0465 |
| *uPH* | Utility coefficient of postoperative hemorrhage | 0.910 | 0.203 | Beta | 0.8986 | 0.0889 |
| *Ug* | Utility coefficient of gastrostomy | 0.916 | 0.209 | Beta | 0.6975 | 0.0640 |
| *Ult* | Utility coefficient of long-term tracheostomy | 0.852 | 0.271 | Beta | 0.6109 | 0.1061 |
| *ues* | Utility coefficient of esophageal stenosis | 0.826 | 0.284 | Beta | 0.6459 | 0.1361 |

| | | | | | | |
|---|---|---|---|---|---|---|
| *uORN* | Utility coefficient of osteoradionecrosis | 0.791 | 0.302 | Beta | 0.6428 | 0.1698 |
| *urem* | Utility coefficient of remission after surgery and adjuvant | 0.980 | 0.099 | Beta | 0.7702 | 0.0346 |
| *uremonlysurg* | Utility coefficient of remission after TORS or TLM alone | 0.957 | 0.151 | Beta | 0.9798 | 0.0200 |
| *ureg* | Utility coefficient of regional recurrence | 0.859 | 0.283 | Beta | 0.4401 | 0.0722 |
| *ulocxrt* | Utility coefficient of local recurrence, RT | 0.771 | 0.302 | Beta | 0.7216 | 0.2143 |
| *uloc* | Utility coefficient of local recurrence, requiring surgery | 0.755 | 0.316 | Beta | 0.6436 | 0.2088 |
| *udist* | Utility coefficient of distant recurrence | 0.213 | 0.336 | Beta | 0.2262 | 0.5106 |
| *upall* | Utility coefficient of palliative care | 0.307 | 0.350 | Beta | 0.1033 | 0.3816 |
